# OCOsense™ smart glasses for analyzing facial expressions using optomyographic sensors

**DOI:** 10.1101/2023.05.12.23289646

**Authors:** James Archer, Ifigeneia Mavridou, Simon Stankoski, M. John Broulidakis, Andrew Cleal, Piotr Walas, Mohsen Fatoorechi, Hristijan Gjoreski, Charles Nduka

**Affiliations:** Emteq Labs, Brighton, East Sussex, BN19SB, United Kingdom; Ss. Cyril and Methodius Unviersity in Skopje, N. Macedonia

## Abstract

This article introduces the Emteq’s OCOsense™ smart glasses equipped with a novel non-contact OCO™ sensor technology for measuring facial muscle activation and expressions based on high resolution tracking of skin movement. We demonstrate that the OCO™ sensor technology based on optomyography is a sensitive and accurate approach for assessing skin movement in 3 dimensions, providing a means for measuring the facial expressions used to assess emotional valence such as smile, frown, and eyebrow raise. We propose that glasses-based optomyography sensing has the potential to herald a paradigm shift in real-world facial expression monitoring, thus enabling real-time emotional analytics with healthcare and research applications.

---

Today, wearable technologies are shaping our everyday experiences. Leveraging familiar form factors, we witness an explosion of smart watches, jewelry-based wearable devices such as rings, earables and eyewear with diverse sensing capabilities [1]. We envision that future wearables cannot only be equipped with multimodal methods for high-accuracy real-time monitoring but could provide biofeedback-informed interventions for a wealth of healthcare and research applications. This will require a robust validation plan in naturalistic test environments.

Compared to other bodily cues, facial expressions are considered the richest source of emotional information [2] (especially of valence; a psychological construct that describes the hedonic component of subjective experience). Facial expression-derived valence is highly dependent on context. Put simply, it is important to know May Published by the IEEE Computer Society what the person is doing to guide affective insights than an expression may convey [3]. We envision that continuous monitoring of facial activation combined with information about the users’ activities can offer insights on the wearer’s emotional state and behavioral changes [4].

In this article we present the OCOsense™ smart glasses equipped with optomyographic (OMG) sensors, which allow real-time monitoring of facial muscle activations. The glasses also include 9-axis Inertial Measurement Unit (IMU) and altimeter for activity and head movement tracking, and speech detection microphones. The OCOsense™ system offers open data access for researchers via its API and a mobile application. In this paper, we present insights on OCO™ sensor performance in tracking skin movement in the laboratory and detecting facial expressions (smile, frown and eyebrow raise).

## RELATED WORK

Studies using methodologies such as affective priming demonstrate that affective changes influence information processing, decision making and interpersonal interaction (e.g. [5], [6]). Therefore, understanding a person’s affect (instinctual emotional response) could be a valuable tool for understanding their behavior. There are several wearable technologies that recognize bodily features as biobehavioral fingerprints. The majority is focused on arousal metrics, using wrist-worn sensors for pulse-rate (e.g., Fitbit^1^, Apple Watch^2^) or electrodermal response (e.g. Empatica^3^). Head-worn devices, (e.g. Nokia’s earable device [1]) are however limited and often in the research phase.

The face and specifically the activation of the zygomaticus major and corrugator muscles has been extensively investigated as a means of recording valence changes [7]. Traditional techniques include surface electromyography (EMG) as the gold-standard measuring muscle contractions, and camera-based tracking [3]. Although monitoring facial activations in this way is commonplace, measuring facial expressions and valence changes naturalistically remains a challenge. There are two reasons for this. The first is the lack of generalizability. A smile can be ironic or even threatening, making the context in which it is elicited critical. Second, although discrete commercial wearables for recording physiological arousal do exist, there is no corresponding discrete method of passively measuring facial expressions reliably, that do not also necessitate a face-directed camera, tethered connection, or sensors placed directly on the skin [8]. In other words, traditional methods are conspicuous by necessity and thus ill-suited for everyday wearable integration.

Recently, novel approaches have been pursued in facial wearables, combining various methods for facial tracking. Amongst those are glasses equipped with capacitive sensors [9], and those with face-mounted cameras [10], which however often suffer limitations with regards to electromagnetic interference and generalized immunity to different environmental conditions such as ambient light. Similarly, electrooculographic (EOG) glasses were developed to detect facial activations and by extension facial expressions [11]. When compared to our sensors approach, the EOG approach is limited by high sensitivity to head movements and low sensitivity to lower-face actions such as smiling. In our proposed solution, head movements do not influence the data to that level because the OCO™ sensors are contact-less. Additionally, due to the sensor type and the locations (around the eyes and on the cheeks) OCO™ sensors can easily distinguish facial expressions such as smiling. Another compelling system utilized photo-reflective sensors on glasses measuring skin proximity (Z-axis) [12]. The Z axis corresponds to the proximity to skin (distance) and the XY axes register movements in the cartesian plane of the skin. Facial muscles produce pulling forces on the skin towards the muscle’s orientation, resulting pronounced protrusions in high intensities [2]. Therefore, proximity alone can be insufficient [13]. We address these issues by introducing the OCO™ sensors capable of measuring skin movements in 3 dimensions, and the potential to detect a wider range of facial activation intensities, such as micro expressions[14]

## OCOSENSE™ GLASSES

The glasses contain seven OCO™ optomyographic sensors [15], a 9-axis IMU, altimeter and dual microphones. The sensors are built into a glasses frame to overlap key facial muscle groups associated with the affective changes (see OCO™ Sensors section) [13]. Figure 1 shows the OCOsense™ glasses, the sensor and the system architecture.

**FIGURE 1.**
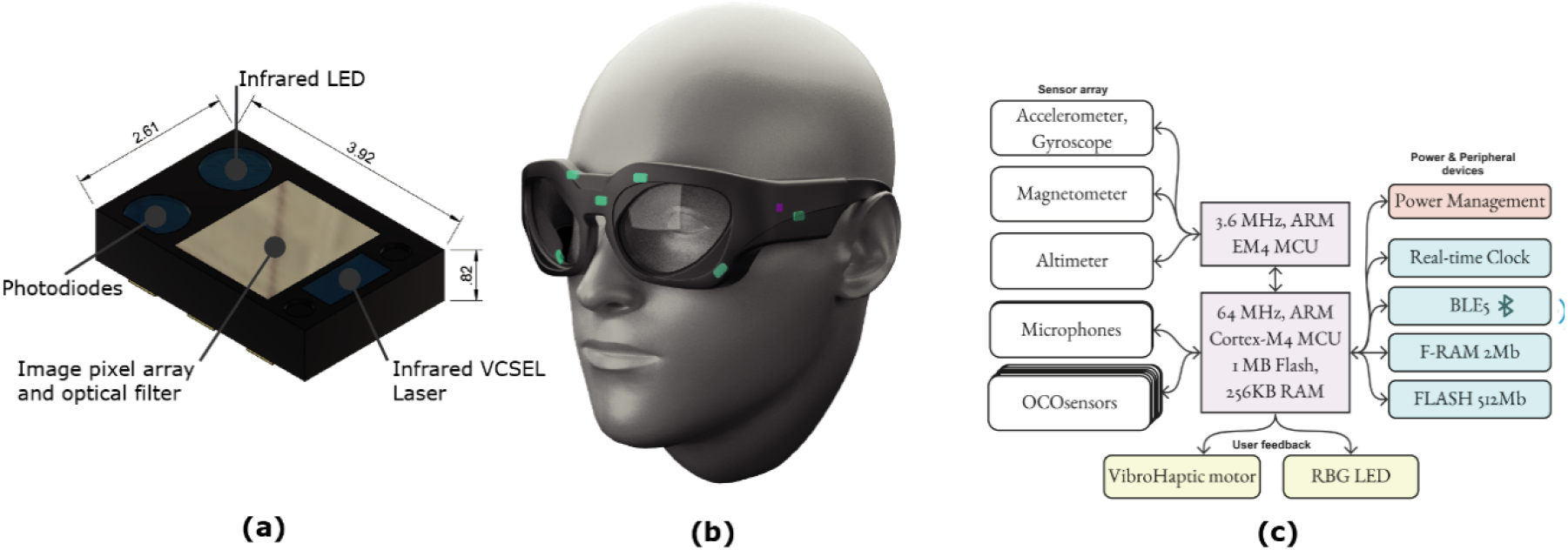
(a) shows a 3D render showing a future OCO™ sensor application specific integrated circuit (ASIC) and its dimensions and fundamental components. (b) shows a 3D-render of the OCO™ glasses on an average head model. The OCO™ sensors (green rectangles), and the IMU (purple rectangle) are highlighted on the model. (c) diagram showing the main electronics components within the OCOsense™ glasses.

All sensors are sampled at 50Hz and streamed via Bluetooth low energy (BLE) to a mobile device. The glasses contain two 220mAh LIPO batteries, which can power the system for 4-6 hours depending on the how active the user is due to the OCOsense™ glasses dynamic framerate power management strategy.

Lastly, the real time clock (RTC) and additional RAM and FLASH memory are used to keep track of the current time and date and store data when the mobile app is not connected to the glasses respectively. A dedicated MCU has also been implemented for onboard real-time activity recognition using machine learning for future research.

### Glasses Design

The OCO™ Sense glasses were designed around statistically generated head models [16] and optimized for comfort and stability whilst worn, thus limiting movement artifacts. Two OCO™ sensors are positioned over the frontalis on the left and right side of the forehead to pick up brow furrow and brow raise movements associated with frown, and eyebrow raise expressions respectively, and two OCO™ sensors are positioned over the zygomaticus major muscles (left and right side of the cheeks) to detect smile expressions. The OCO™ sensor positioned over the procerus (center brow) can be used to help distinguish between frown and brow-raise expressions, only its Z-axis (skin-proximity) measurements are used in this study. Additionally, movement artifacts are detected via two sensors positioned over the left and right temples as shown in Figure 1. The 9-axis IMU and altimeter are positioned in the right temple of the glasses frame.

### The OCO™ Sensor

The OCO™ sensors are a proprietary sensor developed and patented by Emteq Labs (UK patent No.US11,003,899), and use an optical non-contact approach - Optomyography [15], that has advantages over EMG-based systems. EMG electrodes require firm and constant contact with skin to achieve an acceptable signal to noise ratio; the OCO™ sensors are optically based, therefore they do not require skin contact, and can function accurately from 4mm to 30mm from the skin.

OMG has high sensitivity to movement (<4um) and does not require a complex filtering procedure to extract useable signal, which would be computationally expensive and induce detection delays [17]. Furthermore, the output data from the OCO™ sensors are just three 16-bit coordinates positions (X, Y, Z), the sensors are polled at 50Hz by the MCU resulting in a data rate of 2.4kb/s per sensor.

The data rate of all sensors (7 x OCO™ sensors, 9-axis IMU, altimeter and dual speech-detection microphones) is 2.9kB/s, very low compared to camera-based facial expression recognition (FER) systems. An analysis of sensing technologies for FER suggested that active stereoscopy cameras are the most suitable technologies [18]; for example the Intel® RealSense™ Depth Cameras which require a down-stream bandwidth ranging from 13.9MB/s to 146.5MB/s. Additionally, the OCO™ sensors are power-efficient, consuming between 37-68mW depending on the state of the dynamic framerate power management strategy implemented.

The principal operation of the OCO™ sensor and OMG in general, relies on infrared (IR) light sources and sensors to measure skin surface features in the reflected IR light resulting from muscles activating below and around the skin area of interest. The OCO™ sensors use an IR laser, lens-less pixel array, and optical flow logic built into the (ASIC) to calculate muscle-induced movement of the skin.

## OCOTM SENSOR CHARACTERIZATION

### Testing procedure

To validate the OCO™ sensors we developed a specialized 3-axis computer numerical controlled (CNC) movement testing rig, that could accurately move an OCO™ sensor development kit over human skin tissue samples in three dimensions. An example of the CNC test rig itself is shown in Figure 2.

**FIGURE 2.**
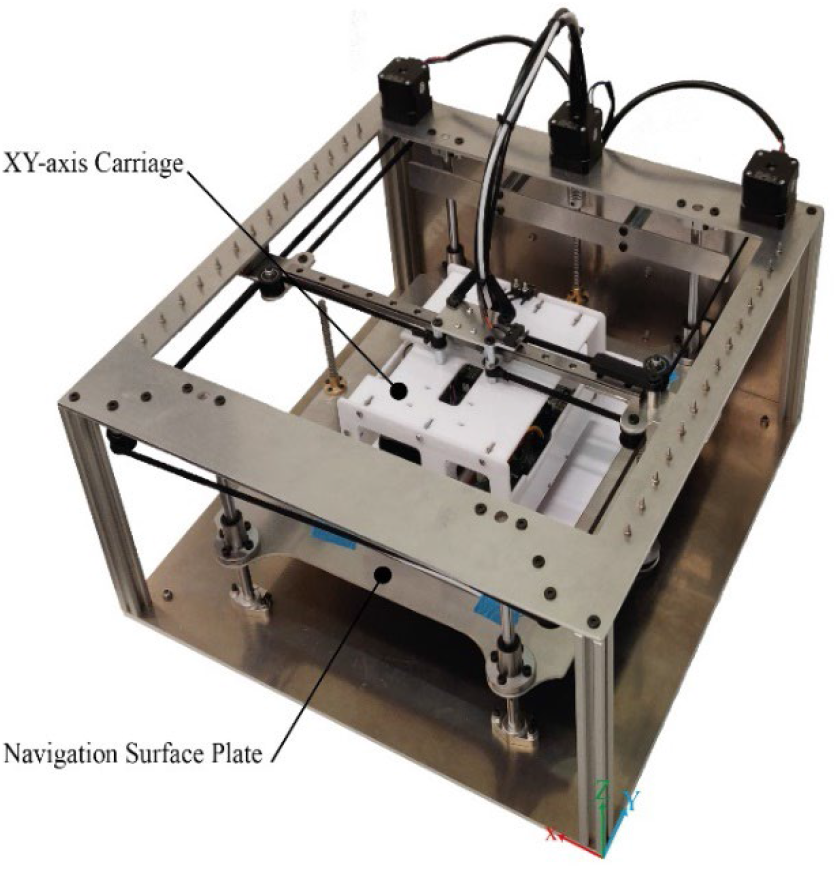
The computer numerical controlled (CNC) skin testing machine developed by Emteq that can move an OCO™ sensor development kit over human skin tissue samples in 3 dimensions. The sensors are mounted in the XY-axis carriage and can be moved in the X&Y-axis, the skin samples can be placed on the navigation surface plate and can then be moved in the Z-axis.

The OCO™ sensor development kits are mounted in the XY-axis carriage and moved using a CoreXY cartesian motion platform^4^. The navigation surface plate moves in the Z-axis and controls the proximity between the sensor and the skin samples. The movement resolution for each axis of the sensor test rig is as follows: X-axis = 12.5 μm, Y-axis = 12.5 μm and Z-axis = 2.5 μm.

To evaluate the movement tracking accuracy of the OCO™ sensors over the skin plane (X&Y-axis) we conducted over 100 movement tests: using circular and linear moment profiles, different velocities (1,000mm/min, 5,000mm/min and 10,000mm/min) and at different proximities from the skin (8mm, 18mm and 28mm). These parameters were chosen based on the maximum facial expression skin movement velocity values determined from literature (∼40mm/s) [19], and by proximity ranges between each of the seven OCO™ sensors and the face that can reasonably occur within our glasses frame when worn correctly (8mm - 28mm).

### OCOTM sensor characterization results

Each time the X&Y axis position is polled by the MCU, the OCO™ sensor outputs the (X, Y) coordinate position relative to the previously polled position. Because the measurements are relative and there is small amount of error associated with each reading, we get an accumulation of error overtime, normally expressed as a drift from the origin as shown in Figure 3.

**FIGURE 3.**
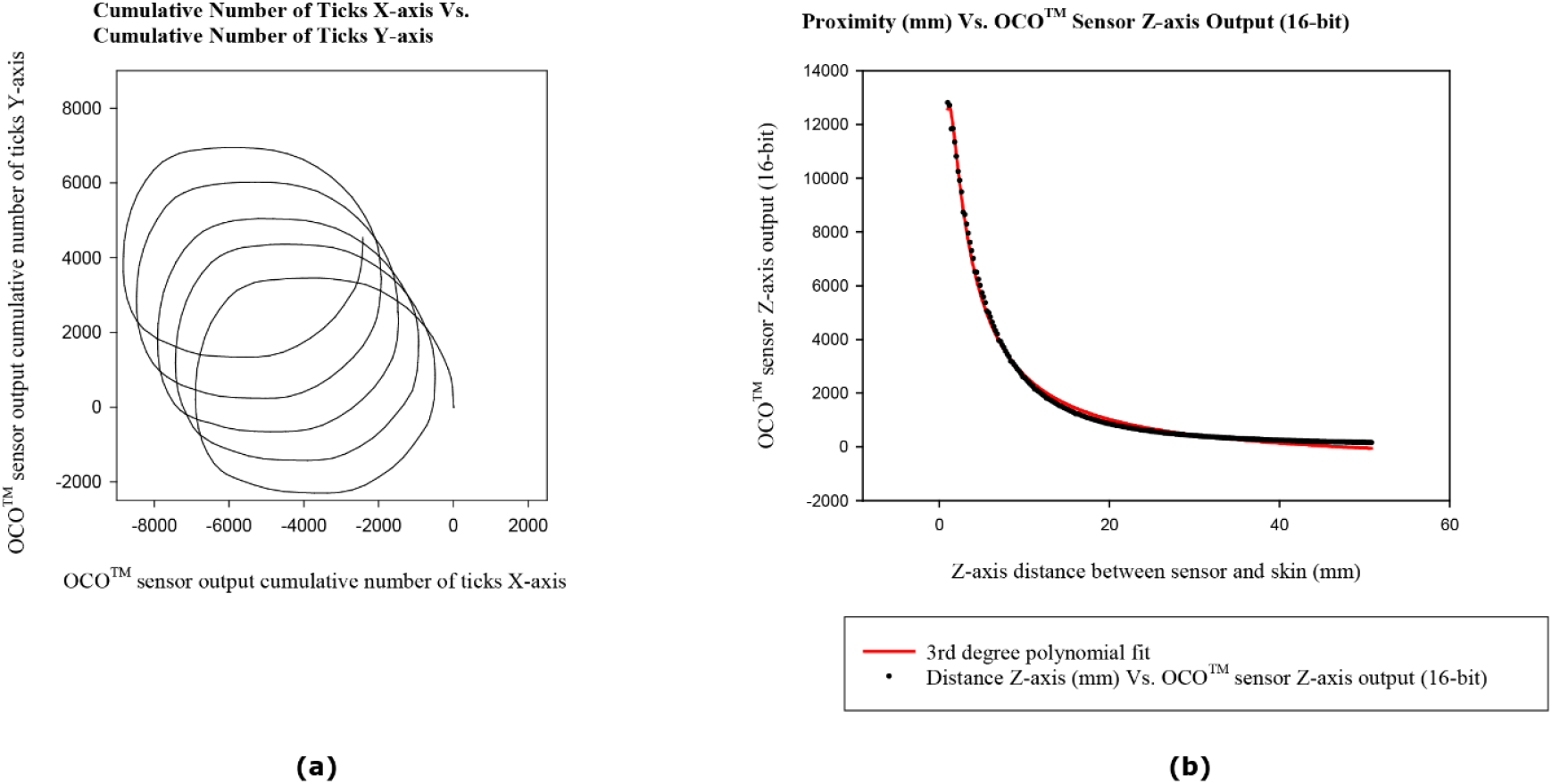
(a) is an example the OCO™ sensor’s X&Y-axis output data, the sensor was moved in a circular motion (15mm radius) over a human skin tissue sample at a velocity of 5,000mm/min at a constant proximity of 18mm, for 5 repetitions. (b) shows the OCO™ sensor’s Z-axis output data, the sensor was moved in the Z-axis from 4mm to 50mm away from the skin sample in 0.2mm increments.

The output units of the OCO™ sensor X&Y-axis are referred to as ticks, the number of ticks per mm depends on the navigation surface and can be influence by skin tone, makeup, perspiration and other refractive influencers. The average ticks per mm value from all the human skin tissue movement tests is 264 ticks/mm, resulting in a skin movement tracking resolution of 3.79μm. Thus, OCO™ sensors could be more accurate compared to alternative camera-based systems attempting use computer vison techniques to track facial landmarks and could allow the OCO™ Sense glasses to pick up micro expressions similar to facial EMG systems.

The error rate of skin tracking in the X&Y-axis is a unitless quantity (ticks/tick, mm/mm, inches/inch, etc.), the average error rate from all our movement tests is 0.027mm/mm. The error-rate is minimal, but it does result in an accumulation of error overtime, which can be problematic if left unchecked.

The OCO™ XY sensing error-rate was constant over the proximity range (8mm to 28mm) and over the movement velocity range (1,000mm/min to 10,000 mm/min) tested, there was no significant difference in error-rate across the board.

A different testing method was required to evaluate the performance of the OCO™ sensor’s Z-axis movement measurement. The sensor was moved in the Z-axis from 4mm to 50mm away from the human skin tissue sample in 0.2mm increments, the resulting sensor output is shown in Figure 3. The relationship between the Z-axis OCO™ sensor output (16-bit) and proximity from the skin (mm) is not linear, but we can fit a 3rd degree polynomial to the data to establish a relationship between them (R^2^ = 0.998).

## FACIAL EXPRESSIONS VALIDATION

To validate the sensors’ ability to detect expression activations on human faces, we performed a study with 18 participants (10 males and 8 females with a mean age of 25.6 ± 3.6 years). The glasses were paired with an iPad running the OCOsense™ mobile application. After simple set-up of the glasses, volunteers were instructed using the mobile app to perform three naturalistic expressions with six repetitions each (six naturalistic smile expressions, followed by six frown expressions and six brow-raise expressions). An example of the data recording is displayed in Figure 4, using data collected from the OCO™ sensors for each of the three axes (X, Y and Z).

**FIGURE 4.**
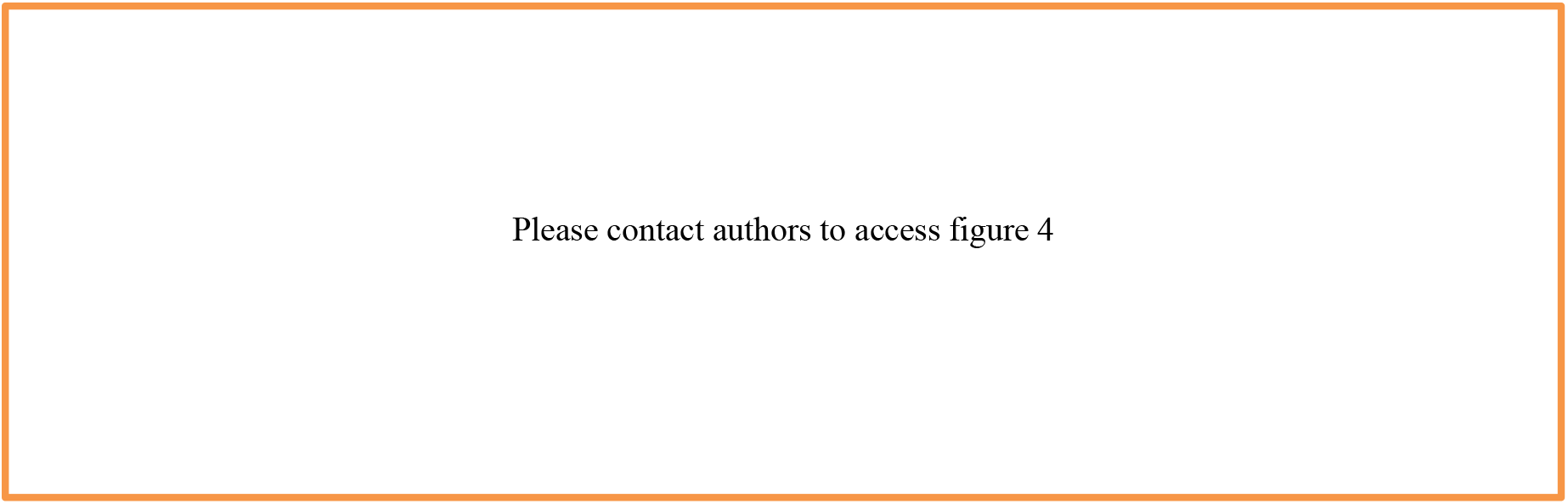
Graphs showing the OCO™ sensor raw data outputs for skin movement on three axes, X, Y and Z. The wearers perform six smile expressions (green regions), then six frown expressions (yellow regions) and six brow-raise expressions (red region).

For the analysis we investigated the relationships between the facial expressions and the values measured by the OCO™ sensors for each expression and direction (X, Y and Z) separately. Firstly, for each subject an average value was calculated for every sensor group (e.g., left and right cheek). The mean normalized amplitude was calculated, for each expression type and for each sensor direction (X, Y, and Z) separately.

Wilcoxon signed-rank (paired) tests with Bonferroni correction were conducted to evaluate whether the OCO™ sensors detected significantly different activations across the sensors and axes. Three pairs of expressions were tested – smile versus frown, smile versus eyebrow raise, and frown versus eyebrow raise. Figure 5 shows that the difference in the normalized amplitude is statistically significant in the majority of the cases (15 out of 21). For each OCOsense™ sensor-direction the results are discussed in the following subsections.

**FIGURE 5.**
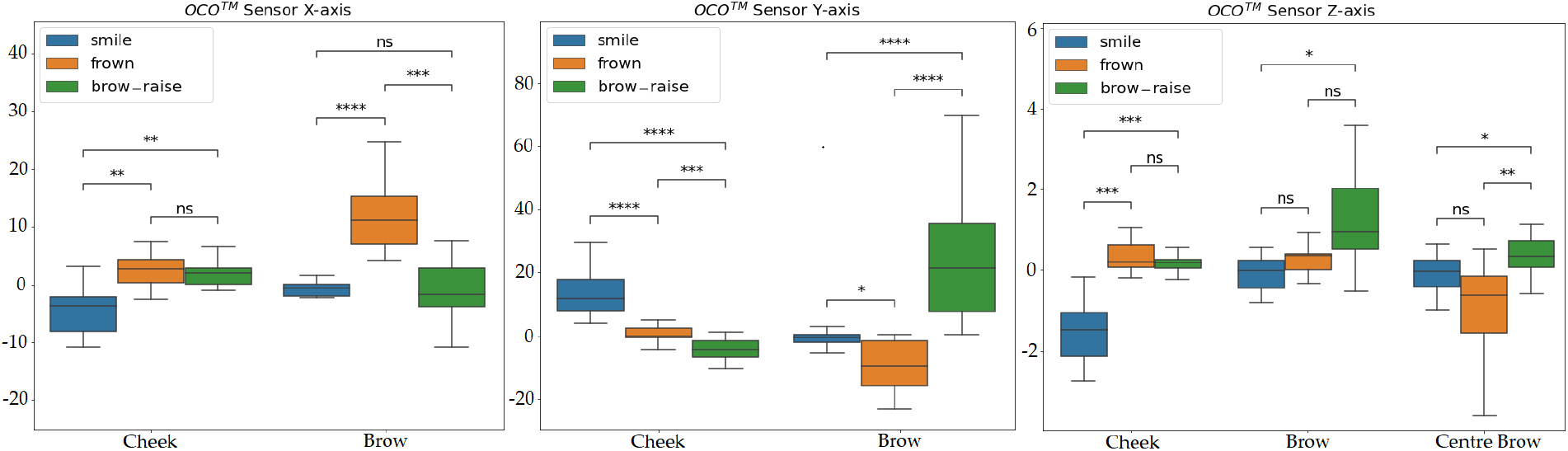
Wilcoxon signed-rank (paired) test with Bonferroni correction for each OCOsense™ sensor activation, for comparison of the following expressions: smile vs frown, smile vs eyebrow raise, and frown vs eyebrow raise. Statistical significance annotations: * if p ∈ [.05, 10−2); ** if p ∈ [10−2, 10−3); *** if p ∈ [10−3, 10−4); and **** if p≥ 10−4.

### THEME/FEATURE/DEPARTMENT

#### OCO− sensor X-axis

The boxplots for the X-axis sensor measurements positioned over the cheeks, show that during smile expression the values of the sensors are significantly different when compared to frown or eyebrow raise expressions. This is expected because the muscles in the cheeks are predominantly activated when smiling. The cheek sensor data during frown and eyebrow raise expressions showed no significant difference.

For the brow sensors, we can see that the biggest activation is measured during the frown expression. The difference between frown and the other two expressions was found to be significant. As expected, the muscle contracting between the brows, the corrugator, is inherently generating skin deformation along the X-axis which is considerably more dominant than in the other two expressions.

#### OCO™ sensor Y-axis

Starting with the cheek sensors, the boxplots show that the largest activation is observed when smiling. This is expected because when smiling the zygomaticus and orbicularis muscles contract by pulling the skin along the Y-axis. As a result, all tests indicate significant difference when comparing smile with the other two expressions. Additionally, we observe a significant difference between frown and brow raise, showing that there is an identifiable upwards cheek movement during the frown expression.

For the brow sensors, it can be clearly seen that the biggest activation on the Y-axis is when performing an eyebrow raise expression. Again, this is expected because the contraction of the frontalis muscle raises the eyebrows and the skin above them. As a result, the statistical tests show significant difference between the data when performing eyebrow raise expression and each of the other two expressions. Furthermore, when frowning, there is an activation of the corrugator muscle that moves the skin downwards. As result all tests were significant.

#### OCO™ sensor Z-axis

For the smile expression the biggest activation is measured by the cheek sensors. This is expected because when smiling the activation of the muscles in the cheeks results in a closer proximity (Z-axis) between the skin and OCO™ sensors in the lower part of the glasses frame. This was also supported by the statistical tests. The cheek activation during frown and eyebrow raise was overall low (higher proximity distance) showing no significant difference.

The boxplots of the OCO™ brow sensors showed increased distance between the sensors and the skin during the brow raise expression. This could be caused by the skin stretching thus reducing its volume. When comparing to the other expressions, the proximity value was different only between the eyebrow raise and the smile expression, where the distance was slightly lowered.

The biggest overall activation on the central brow sensor was detected during frowning. This is expected because frowning activates the corrugator muscle which results in protruding the skin between the brows. Therefore, the distance from the sensor to the skin is reduced. Contrary to frowning, when raising the eyebrows, the skin between the brows stretches, decreasing skin-distance. The statistical tests also support this difference. However, when comparing the frowning and smile expression no significant difference was found. This is not completely expected because usually when we are smiling no activation occurs between the brows. Nonetheless, based on research done in this field [20], corrugator activity can be seen for some people when smiling.

## CONCLUSION

In this paper we presented the OCOsense™ smart glasses capable of continuous and accurate measurement of facial skin movement caused by muscle activation. The device features novel OCO™ sensors, OMG-based, which measure skin movement in three dimensions over key facial muscles.

We performed CNC movement tests, which showed that the OCO™ sensor skin tracking resolution in the XY-plane is less 4 μm. Also, the error rate of the skin tracking along the XY-plane was consistently 0.027mm/mm over the sensor-skin proximity range (8mm-28mm), and over a velocity range synonymous to facial expressions (1,000mm/min to 10,000mm/min). Furthermore, we validated accurate measurement of proximity to the skin (Z-axis) from 4-50mm in 0.2mm increments. The combined rate required for seven OCO™ sensors (2.4kb/s per sensor) is significantly lower compared to FER camera-based systems and thus less computationally expensive.

Further tests with participants confirmed that using the OCO™ data we can distinguish three facial expressions: smile, frown and brow-raise. The experiments indicated statistical differences in the sensor data when the 3 expressions are performed, i.e., specific groups of sensors revealed elevated activations during expressions, e.g., cheek sensors during smile, center-brow sensor during frown and brow sensors during brow-raise. Also, the results showed that by using only X-axis sensor data or only the Z-axis, we cannot differentiate between all pairs of expressions. However, the Y-axis data analysis shows significant difference for all expressions. The combination of all axes could provide a robust method for accurate expressions detection [13].

Traditional methods for facial movement tracking are often conspicuous and difficult to use outside the lab; for example methods such as EMG and EOG require skin contact and are susceptible to motion artefacts, and camera-based methods directed at the face can impair the user’s interaction with the environment. The OCOsense™ platform presented in this paper was designed to address limitations in previous wearable sensing eyewear and inform future research in the area. Further validation in longitudinal and naturalistic settings will be performed as part of our future work. With this system we envision informing solutions for viable non-invasive and inconspicuous wearable facial-activation measurement, which could be applied outside the laboratory, on a variety of applications and settings including augmented reality.

## LIMITATIONS AND FUTURE WORK

One limitation of the glasses is that the X&Y-axis skin measurements are relative to each other, which results in cumulative error over time when calculating the XY-plane coordinates. This can be potentially addressed by recalibrating over time, i.e., resetting the drift in the XY-axis during periods of repose.

In general, the OCOsense™ glasses work well in varying ambient light conditions due to the presence of IR pass filters and ambient light compensation circuitry. However, in future studies we aim to quantify the limitations of the device in extreme lighting conditions. Furthermore, other environmental factors could affect sensor accuracy such as skin surface changes including perspiration, make-up, hair which will be investigated.

We are currently conducting larger-scale data collection and clinical evaluation studies for measuring facial expressions, activities and emotional valence. These will allow us to develop machine-learning algorithms to recognize various expressions, activities of daily-living, and to validate how well the algorithms generalize over a large-scale population. These studies will also provide insights into the effect of population difference on product fit and emotional response. The glasses’ fit on the wearer is very important i.e., the data will differ on different people’s faces. These require the development of calibration and personalization algorithms so that the models are adjusted to the wearer’s face characteristics.

Future work could involve utilizing the integrated IMU and altimeter for recognizing activities of daily living [14]. Also, the dual microphone can be used for speech detection capabilities and vocal prosody analysis. All this could enable future research and development to create advanced machine learning algorithms for insights into context-aware valence, whilst supporting future ubiquitous applications in virtual and physical product evaluation, research, healthcare, and wellbeing.

## Data Availability

All data produced in the present study are available upon reasonable request to the authors

## ACKNOWLEDGMENTS

Development of the OCOsense™ system was part-funded by grants awarded by the NIHR (NIHR PDA award; ref: II-LA-0814-20008) and Innovate UK (Investment accelerator. ref: 105207).

**James Archer**, leads hardware development for the OCOsenseTM platform at Emteq Labs. He received his PhD from the Sensor Technology Research Centre University of Sussex. Contact him at.

**Ifigeneia Mavridou**, is the Lead Affect Engineer at Emteq Labs. Mavridou received her EngD on affect recognition from physiological signals. Honorary Research Fellow at Bournemouth University and Sussex University. Contact her at.

**Simon Stankoski** is Data Scientist at Emteq Labs. He finished his master studies at Jožef Stefan International Postgraduate School in Ljubljana, Slovenia. Contact him at

**John Broulidakis**, Head of Clinical Research at Emteq Labs, and is responsible for validating OCOsense™. He received his PhD from the University of Southampton. Contact him at

**Andrew Cleal**, is a Lead Software Developer at Emteq Labs in Brighton, UK. Contact him at.

**Piotr Walas** is a Firmware Engineer at Emteq Labs in Brighton, UK. Contact him at.

**Mohsen Fatoorechi** is a Lead Hardware Engineer at Emteq Labs in Brighton, UK. He received his PhD from the Sensor Technology Research Centre University of Sussex. Contact him at.

**Hristijan Gjoreski**, is Associate Professor at Ss. Cyril and Methodius University in Skopje, N. Macedonia and is the

Lead Data Scientist at Emteq Labs in Brighton, UK. Contact him at

**Charles Nduka**, is the CEO and Chief Technologist Emteq Labs in Brighton, UK. Contact him at

## Footnotes

1 https://www.fitbit.com/global/us/technology

2 https://www.apple.com/uk/watch/

3 https://www.empatica.com/en-eu/research/e4/

4 https://corexy.com/

